# Cross-Attention Enables Context-Aware Multimodal Skin Lesion Diagnosis

**DOI:** 10.64898/2026.03.10.26348046

**Authors:** Krishna Mridha, Humayera Islam

## Abstract

Clinical diagnosis of skin lesions integrates visual dermoscopic features with patient context such as age, skin type, and lesion characteristics. However, most artificial intelligence systems for dermoscopic analysis rely solely on image data and ignore structured clinical metadata. We developed a multimodal deep learning framework that combines dermoscopic images with patient metadata and evaluated whether cross-attention mechanisms better capture contextual interactions than conventional fusion strategies. Using 1,568 lesions from the PAD-UFES-20 dataset (69% malignant) with associated metadata (age, sex, Fitzpatrick skin type, anatomical site, and lesion diameter), we compared four models: metadata-only logistic regression, image-only ResNet18, late fusion via feature concatenation, and cross-attention-based fusion. The image-only model achieved strong discrimination (AUC 0.9776), while late fusion slightly reduced performance (AUC 0.9717). The proposed cross-attention model achieved the best overall results (AUC 0.9818, AUPRC 0.9924) with improved calibration (ECE 0.0379). These findings suggest that attention-based multimodal learning enables more effective integration of patient context for automated skin lesion diagnosis.

## Introduction

Skin cancer is one of the most common malignancies worldwide, and early detection is critical for reducing morbidity and mortality. In clinical practice, dermatologic diagnosis is inherently contextual: clinicians evaluate visual characteristics of lesions together with patient-specific factors such as age, anatomical location, and skin phenotype. Morphologic features including asymmetry, border irregularity, color variation, and lesion diameter are interpreted in light of these contextual attributes to estimate malignancy risk.^1^ Formally, this task can be expressed as estimating the conditional probability *P* (*y* = 1 |*I*, **m**), where *I* ∈ ℝ^*H×W ×*3^ denotes the dermoscopic image and **m** represents structured clinical metadata. This formulation highlights the intrinsically multimodal nature of dermatologic reasoning.

Deep learning has substantially improved automated skin lesion analysis. Convolutional neural networks have achieved dermatologist-level performance in image classification tasks,^2^ with subsequent studies demonstrating strong diagnostic accuracy across benchmark datasets.^3,4,5^ Residual architectures such as ResNet^6^ have become standard backbones for dermoscopic analysis, while transformer-based models^7^ have further advanced visual representation learning. Despite these advances, most existing systems treat diagnosis as a purely visual task and estimate *P* (*y* = 1 |*I*) without incorporating patient context. This limitation is particularly important in dermatology, where lesion interpretation depends on demographic and phenotypic variability, and disparities in model performance across skin tones have been reported.^8^

Multimodal learning provides a natural framework for integrating medical images with structured clinical data. Prior work has combined image features with tabular metadata using late-fusion strategies that concatenate independently learned representations before classification.^9^ Similar approaches have been explored across biomedical AI applications.^10,11,12^ However, these methods typically allow interaction between modalities only at the final prediction stage, limiting the ability of clinical context to influence visual feature interpretation.^13,14,15^

Attention mechanisms offer a principled approach to modeling such cross-modal interactions. Introduced in neural machine translation^16^ and generalized in the Transformer architecture,^17^ attention enables one representation to dynamically weight another based on contextual relevance. Conceptually, this aligns with clinical reasoning, where contextual patient information informs how visual findings are interpreted rather than being appended after image analysis.

In this work, we develop a context-aware multimodal framework for skin lesion classification using metadata-guided cross-attention. Dermoscopic images are encoded using a Vision Transformer that produces spatial token representations, while structured clinical variables are represented as learnable metadata tokens. Cross-attention enables metadata tokens to query image tokens, allowing patient context to guide attention over visual features prior to classification. We evaluate this approach on the PAD-UFES-20 dataset against three baseline strategies: metadata-only logistic regression, image-only ResNet18, and conventional late-fusion multimodal learning. Results show that while simple concatenation provides limited benefit, cross-attention enables more effective multimodal integration and improves both discrimination and calibration.

### Contributions

The contributions of this work are threefold. First, we propose a context-aware multimodal architecture that integrates dermoscopic images and structured clinical metadata through metadata-guided cross-attention, allowing patient context to influence spatial visual representations. Second, we systematically compare metadata-only, image-only, late-fusion, and cross-attention multimodal models to quantify the impact of different integration strategies. Third, we provide interpretability analyses, including permutation-based feature importance and case-based examination, to characterize how clinical variables influence model predictions.

## Methods

### Dataset

We conducted a retrospective study using the PAD-UFES-20 dataset,^18^ a clinically annotated collection of smartphone-acquired dermoscopic images collected in Brazilian dermatology clinics. Each case includes a dermoscopic image together with structured clinical metadata.

After filtering for samples with available images and metadata, the final dataset consisted of *N* = 1,568 lesions. The dataset provides demographic variables, including age (continuous) and sex (binary). Lesion-specific characteristics include the lesion diameter measured in millimeters (continuous) and the anatomical site of the lesion (five categorical locations). In addition, patient phenotype information is represented by the Fitzpatrick skin type (I–VI). The ground truth label for each case is derived from histopathological diagnosis and indicates whether the lesion is benign or malignant. Although PAD-UFES-20 contains six diagnostic categories (BCC, SCC, ACK, MEL, NEV, and SEK), a binary malignancy label derived from histopathology is also provided. In this study we directly used this binary label (benign versus malignant) as the prediction target. The dataset contains 1,089 malignant lesions (69%) and 479 benign lesions (31%).

### Patient-Level Data Splitting

To prevent information leakage, dataset partitioning was performed at the patient level using `GroupShuffleSplit`. All lesions belonging to the same patient were assigned to the same partition. The dataset was divided into 80% training and 20% testing sets, with a validation subset further created from the training set using a second patient-wise split.

### Data Preprocessing

Dermoscopic images were resized to 224 ×224 pixels and normalized using ImageNet statistics. During training, data augmentation was applied to improve robustness to variations in lesion appearance. Augmentations included random resized cropping, horizontal flipping, mild rotations, and small color perturbations. Validation and test images were processed deterministically without augmentation.

Clinical metadata consisted of numerical and categorical variables. Numerical features (age and lesion diameter) were standardized using the affine transformation computed from the training set. Missing values were imputed using the training-set median. Categorical variables included sex, Fitzpatrick skin type, and anatomical site. These preprocessing steps were applied consistently across all modeling approaches. However, the final feature representation differed across models, as described in the modeling sections.

### Modeling Approaches

To systematically evaluate the predictive contribution of dermoscopic images, clinical metadata, and multimodal integration, we implemented four modeling strategies with increasing representational capacity: (1) metadata-only prediction, (2) image-only prediction, (3) multimodal late fusion, and (4) the proposed metadata-guided cross-attention multimodal architecture.

### Metadata-Only Mode

The metadata-only baseline predicts malignancy risk using structured clinical variables without incorporating image information. After preprocessing, numerical variables (age and lesion diameter) were standardized using statistics computed from the training set, and categorical variables (sex, Fitzpatrick skin type, and anatomical site) were encoded using one-hot representations.

The resulting metadata feature vector is

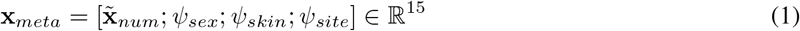

where 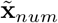 denotes standardized numerical variables and *ψ*(·) represents one-hot encodings for categorical features. Malignancy probability was estimated using logistic regression

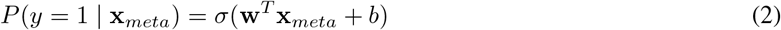

where *σ*(·) denotes the logistic sigmoid function and (**w**, *b*) are learnable parameters. This model serves as a baseline for assessing the predictive value of clinical metadata alone.

### Image-Only Model

The image-only model predicts malignancy directly from dermoscopic images using a convolutional neural network based on the ResNet18 architecture. Images were resized to 224 ×224 pixels and provided as input to the network.

Let

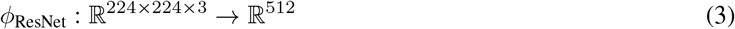

denote the ResNet18 encoder excluding the final classification layer. For an input image *I*, the network produces a visual feature representation

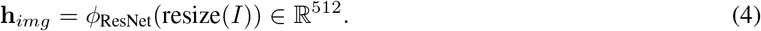

The malignancy probability is then computed through a logistic classification layer

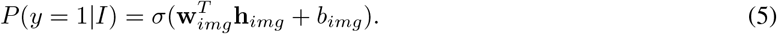

ResNet architectures learn hierarchical visual features through residual learning. Each residual block updates the hidden representation according to

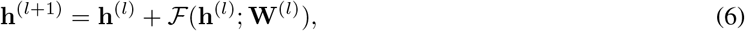

where ℱ (·) represents stacked convolutional layers followed by batch normalization and ReLU activation functions. These residual connections enable stable training of deep networks by facilitating gradient propagation across layers.

#### Late Fusion Multimodal Model

The late fusion model integrates image features and clinical metadata through feature-level concatenation. Visual representations are extracted using the ResNet18 encoder described above, producing

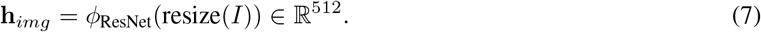

Clinical metadata are represented by the encoded feature vector

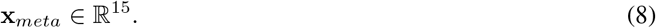

The two modalities are combined by concatenating their feature representations

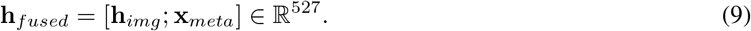

The fused representation is provided to a logistic classifier that estimates malignancy probability

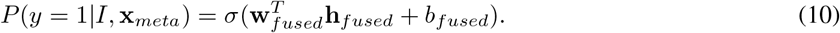

This late fusion approach allows both modalities to contribute to prediction while maintaining independent feature extraction pipelines. However, interactions between clinical variables and spatial image features are modeled only implicitly through the final classifier.

### Proposed Cross-Attention Multimodal Model

To jointly model dermoscopic images and structured clinical variables, we developed a multimodal architecture that integrates visual and metadata representations through crossattention.

Dermoscopic images were encoded using a pretrained Vision Transformer (ViT-B/16). Instead of using only the global class token, the full sequence of transformer tokens, including the class token and patch tokens, was retained in order to preserve spatial information across the lesion. Let

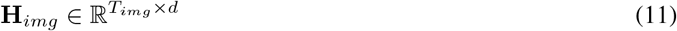

denote the sequence of visual tokens extracted by the transformer encoder, where *T*_*img*_ is the number of tokens and *d* is the hidden dimension.

Clinical metadata were represented as a sequence of learnable metadata tokens. Each categorical variable (sex, Fitzpatrick skin type, and anatomical site) was embedded through a dedicated embedding layer and projected into the same latent space as the visual tokens. Numerical variables (age and lesion diameter) were normalized and concatenated with binary missingness indicators before projection into a numerical metadata token. These transformations produced a metadata token sequence

#### Algorithm 1

Forward Pass of the Metadata-Guided Cross-Attention Model

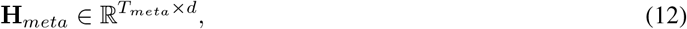

where *T*_*meta*_ denotes the number of metadata tokens representing patient context.

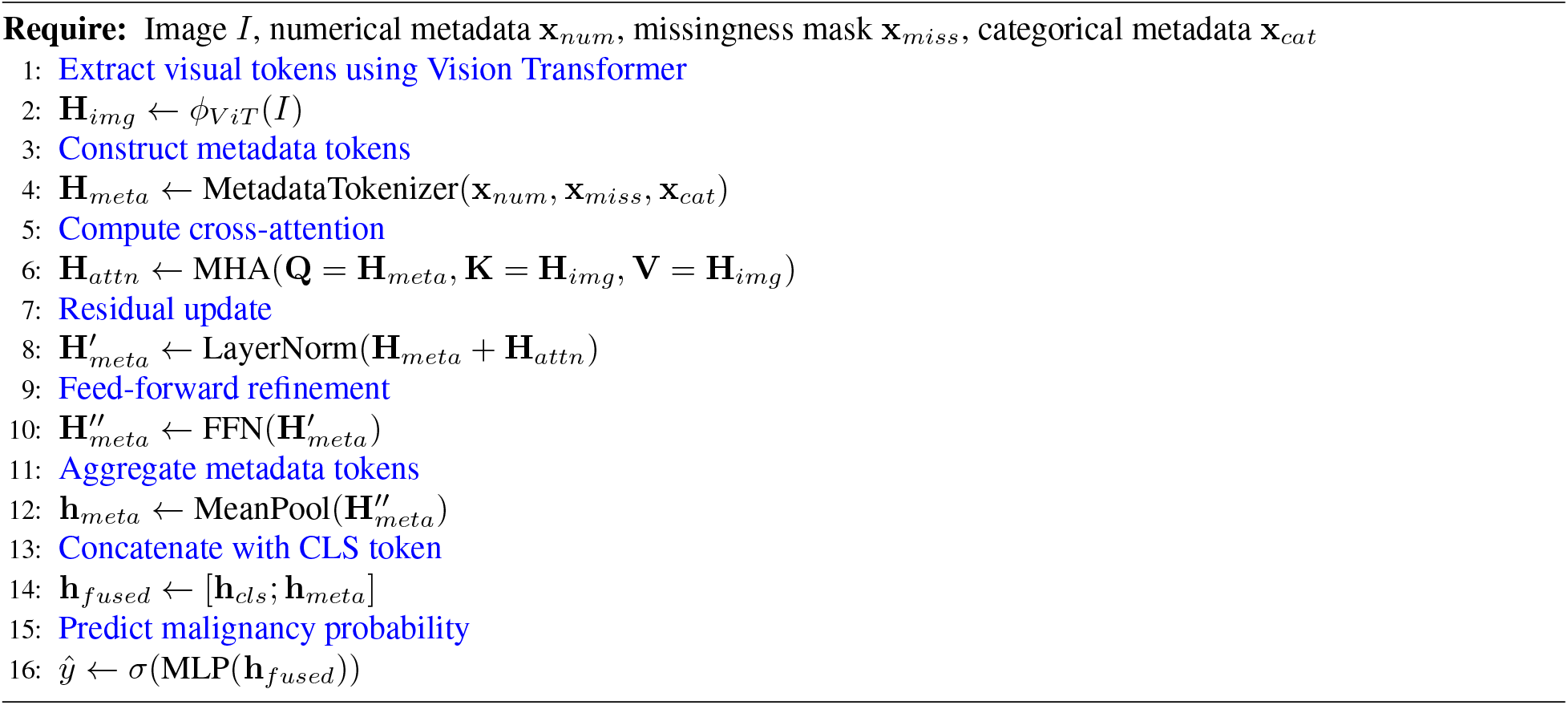

Multimodal fusion was performed using multi-head cross-attention in which metadata tokens act as queries and visual tokens serve as keys and values. The attention operation is defined as

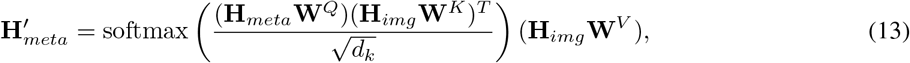

where **W**^*Q*^, **W**^*K*^, **W**^*V*^ are learned projection matrices and *d*_*k*_ denotes the key dimension.

This formulation allows each metadata token to selectively attend to spatial regions of the lesion representation. As a result, patient-specific clinical information can dynamically guide which visual patterns are emphasized during prediction.

The attention output was refined through residual connections, layer normalization, and a feed-forward transformation. The resulting metadata tokens were aggregated using mean pooling to produce a metadata-informed lesion representation

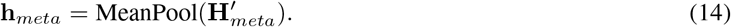

This representation was concatenated with the Vision Transformer class token **h**_*cls*_, which captures the global visual context

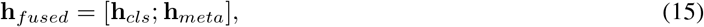

and the fused representation was passed through a multilayer perceptron to estimate the probability of malignancy. Algorithm 1 summarizes the forward computation of the proposed multimodal model.

### Model Training and Evaluation

Training was designed to ensure robust generalization while addressing class imbalance. To mitigate the imbalance between malignant and benign lesions, balanced sampling was implemented using a weighted random sampler that increased the frequency of benign samples during training.

Optimization was performed using the AdamW optimizer with learning rate 3 ×10^*−*4^ and weight decay 10^*−*4^. For the proposed multimodal architecture, the pretrained Vision Transformer backbone was frozen and only the metadata encoder, cross-attention layers, and classification head were optimized. This strategy stabilizes training and reduces the risk of overfitting when the number of labeled samples is limited relative to the capacity of the backbone network.

Binary cross-entropy with logits was used as the training loss. To improve probability calibration and reduce overconfident predictions, label smoothing was applied to the training targets. The learning rate was adaptively adjusted using a ReduceLROnPlateau scheduler, and early stopping was applied based on validation precision–recall AUC.

Model performance was evaluated on the held-out test set using the area under the receiver operating characteristic curve (ROC AUC) and the area under the precision–recall curve (PR AUC). Calibration performance was assessed using expected calibration error (ECE) and Brier score measured the accuracy of predictions (value 0 denotes perfect accuracy). To assess statistical significance between competing models, paired bootstrap resampling with *B* = 2000 iterations was performed. For each bootstrap sample, AUC values were recomputed and differences between models were used to estimate confidence intervals and two-sided *p*-values.

### Model Explainability and Post-hoc Analysis

To better understand how the multimodal model integrates visual and clinical information, we performed post-hoc interpretability analyses on the trained cross-attention model.

First, we conducted a case-based analysis by examining representative examples of correct and incorrect predictions. For each case, the dermoscopic image, predicted malignancy probability, associated clinical metadata, and cross-attention maps were jointly inspected. This qualitative analysis allowed us to assess how the model combines lesion appearance with patient context during prediction and to identify patterns associated with correct classifications and errors.

Second, we performed a permutation-based feature importance analysis to quantify the contribution of individual metadata variables. In this analysis, the trained model was kept fixed while the values of one clinical variable at a time were randomly permuted across the test set, thereby disrupting its association with the correct sample while preserving its distribution. Model performance was then re-evaluated using ROC AUC and PR AUC, and the change relative to the original full-model performance was recorded. Larger performance drops indicate greater reliance of the model on that feature. We also evaluated a no-metadata condition in which all clinical variables were removed, providing an image-only reference within the same trained framework.

### Theoretical Analysis of Cross-Attention

#### Cross-Attention as Kernel Regression

The cross-attention mechanism can be interpreted through the perspective of kernel regression. Let **z**_*meta*_ denote a metadata token and 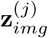 denote the *j*-th visual token extracted from the transformer encoder. The compatibility between a metadata token and an image token can be expressed as

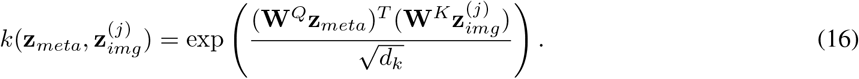

The normalized attention weight for each image token is

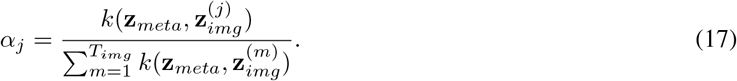

The resulting metadata-conditioned visual representation is therefore

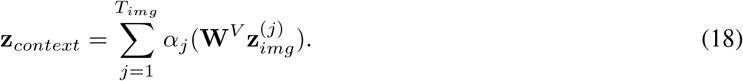

From this perspective, cross-attention performs a kernel-weighted aggregation of visual tokens, where the similarity between transformed metadata and image representations determines the effective contribution of each spatial region.

#### Gradient Flow Analysis

Cross-attention also provides a direct pathway through which metadata influences the final prediction. Let *ŷ* denote the predicted malignancy probability and **h**_*fused*_ the fused multimodal representation used for classification. The sensitivity of the prediction with respect to metadata tokens can be expressed as

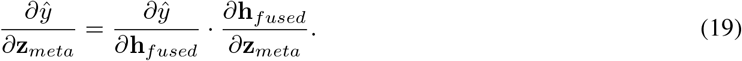

Because metadata tokens participate directly in the attention computation, gradients propagate from the prediction layer through the cross-attention module to the metadata encoder. This mechanism enables end-to-end learning of metadata-conditioned visual representations, allowing the model to dynamically adjust spatial attention patterns based on patient-specific clinical context.

## Results

### Model Performance

Table 1 summarizes the performance of the four models. The image-only model already provides strong discrimination (AUC = 0.9776), whereas naive late fusion slightly degrades performance (AUC = 0.9717), suggesting that simple concatenation of metadata may introduce noise rather than useful context. In contrast, the cross-attention model achieves the best discrimination (AUC = 0.9818, AUPRC = 0.9924) and the strongest calibration (ECE = 0.0379; Brier = 0.0323), indicating that structured integration of clinical metadata improves both predictive accuracy and reliability of probability estimates.

**Table 1:**
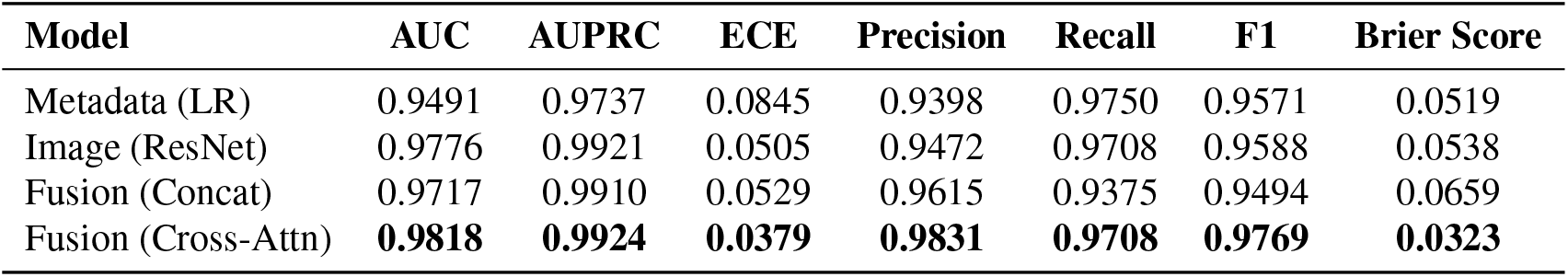
Clinical performance comparison across models. The cross-attention fusion model achieves the highest AUC, AUPRC, and F1-score, and the lowest calibration error (ECE).

### Training Behavior and Predictive Performance

Figure 2 illustrates the training dynamics and predictive performance of the evaluated models. The proposed cross-attention model converges smoothly during training, with training and validation losses decreasing in parallel, indicating stable optimization and limited overfitting (Fig. 2a). Receiver operating characteristic curves (Fig. 2b) further show that the cross-attention model achieves the strongest discrimination among all models. Similarly, precision–recall curves (Fig. 2c) demonstrate improved performance under class imbalance, with the cross-attention model maintaining higher precision across a wide range of recall values.

**Figure 1:**
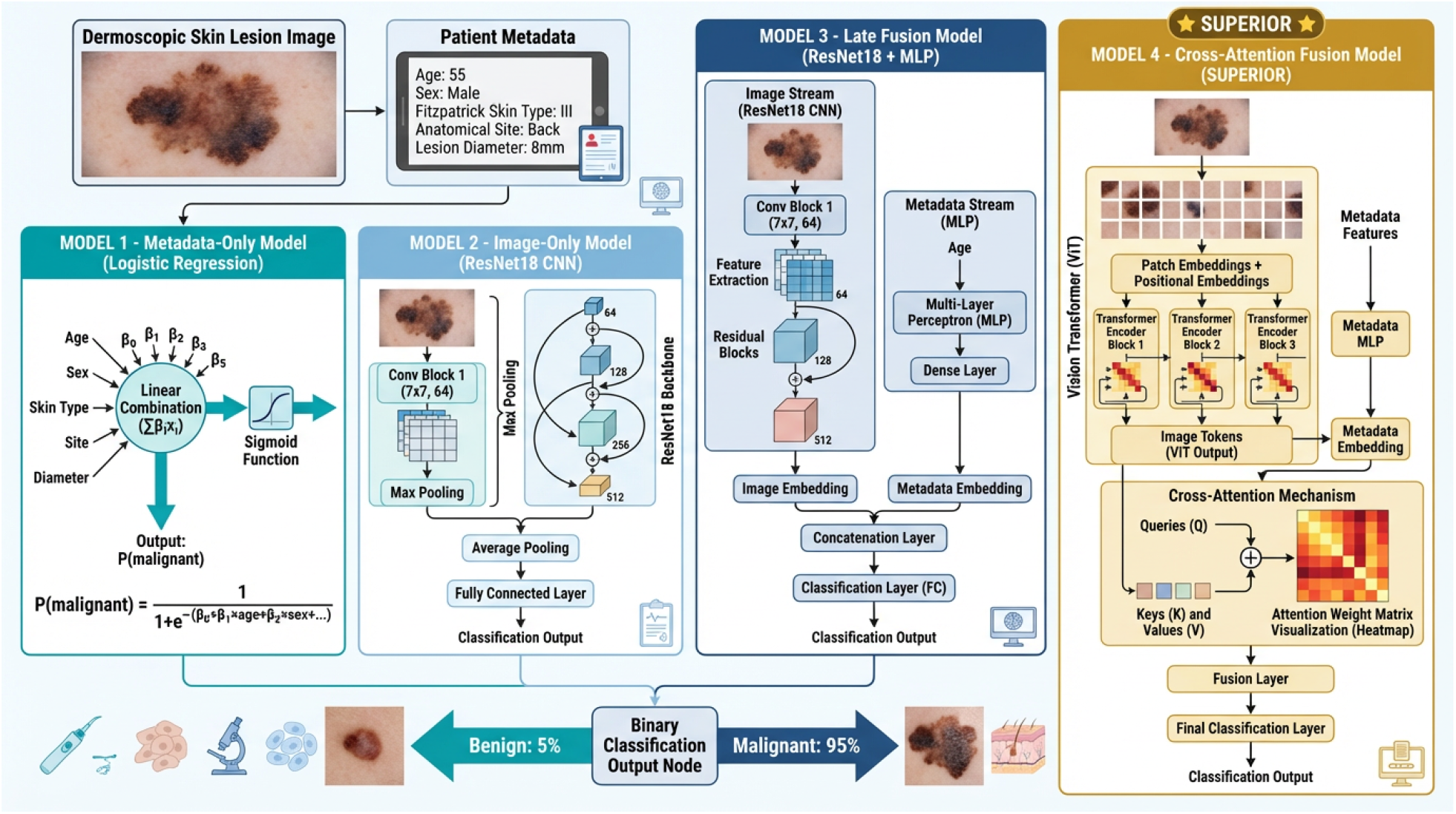
Overview of the multimodal framework for context-aware skin lesion classification. The model integrates dermoscopic images with structured clinical metadata (age, sex, Fitzpatrick skin type, anatomical site, and lesion diameter). Four modeling strategies are evaluated: (1) metadata-only logistic regression, (2) image-only convolutional neural network (ResNet18), (3) multimodal late fusion through feature concatenation, and (4) the proposed cross-attention multimodal architecture. In the proposed model, metadata tokens attend to visual tokens extracted by a Vision Transformer, enabling metadata-guided interpretation of lesion morphology prior to classification.

**Figure 2:**
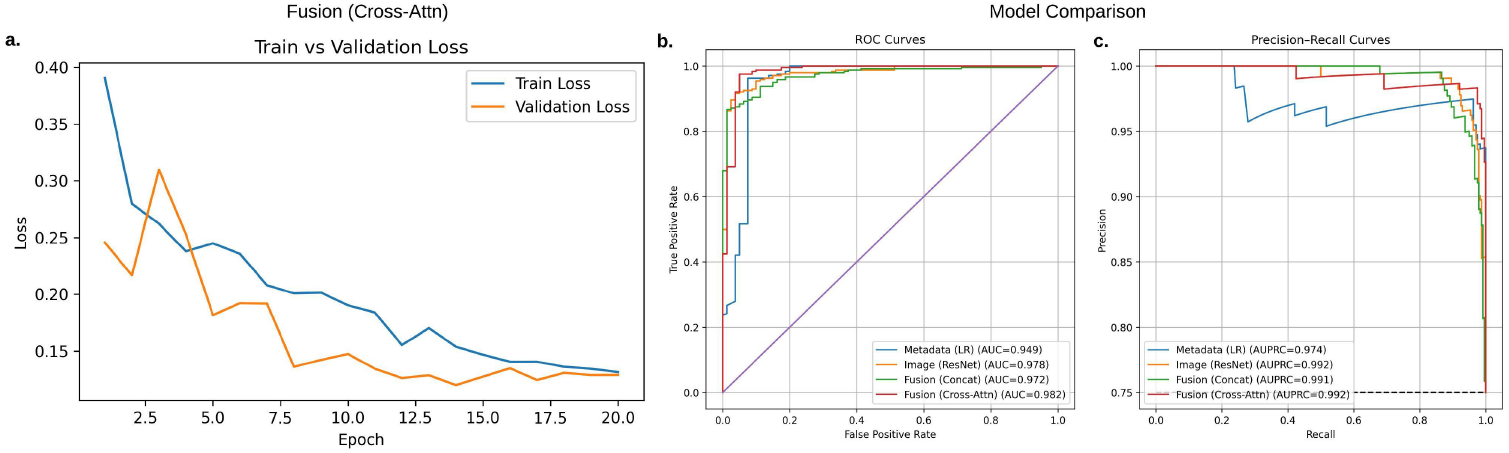
Training dynamics and comparative performance of multimodal skin lesion classification models. (**a**) Training and validation loss curves for the cross-attention model showing stable convergence. (**b**) ROC curves comparing metadata-only logistic regression, image-only ResNet, late fusion, and cross-attention fusion. (**c**) Precision–recall curves demonstrating improved performance of the cross-attention model under class imbalance.

#### Error Patterns

Figure 3 presents normalized confusion matrices for the four models. The cross-attention model achieves the highest specificity, reducing false positives for benign lesions while maintaining a low false negative rate for malignant cases. This balance between sensitivity and specificity suggests that structured multimodal fusion improves diagnostic precision without sacrificing the ability to detect malignant lesions.

**Figure 3:**
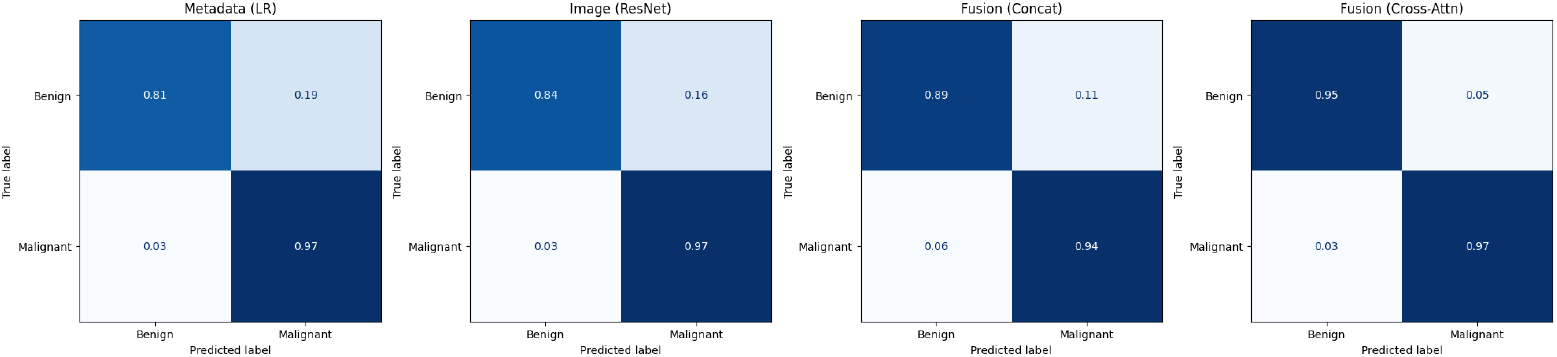
Normalized confusion matrices for the four evaluated models. The cross-attention model reduces false positives for benign lesions while maintaining a low false negative rate for malignant cases, indicating improved diagnostic precision.

#### Statistical Comparison of Fusion Strategies

Table 2 reports paired bootstrap comparisons of model performance. Although the cross-attention model achieves the highest AUC overall, the improvement over the image-only baseline is modest and not statistically significant (ΔAUC = 0.0044, *p* = 0.687). Similarly, late fusion does not significantly improve over the image-only model and shows a slightly negative point estimate (ΔAUC = −0.0059, *p* = 0.167). These results suggest that while structured multimodal fusion can yield the best empirical performance, gains over strong visual models may be limited in relatively small datasets.

**Table 2:** Paired bootstrap comparison of model performance (*B* = 2000 resamples).

| Comparison | $\Delta\text{AUC}$ | % Gain | 95% CI | p-value |
| --- | --- | --- | --- | --- |
| Fusion (Concat) vs Metadata | +0.0224 | 2.36% | (-0.0137, 0.0651) | 0.2620 |
| Fusion (Concat) vs Image | -0.0059 | -0.60% | (-0.0142, 0.0027) | 0.1670 |
| Cross-Attn vs Fusion (Concat) | +0.0102 | 1.05% | (-0.0105, 0.0304) | 0.3190 |
| Cross-Attn vs Image | +0.0044 | 0.45% | (-0.0192, 0.0257) | 0.6870 |

#### Attention-Based Interpretability

Figure 4 illustrates representative cross-attention maps for correctly and incorrectly classified samples. For correctly classified malignant lesions, attention consistently focuses on irregular pigmentation and structural features within the lesion. True negative cases show attention concentrated on benign patterns or surrounding skin regions. In contrast, misclassified cases exhibit more diffuse or misplaced attention, suggesting that ambiguous visual patterns or limited metadata context may contribute to incorrect predictions. These observations indicate that the cross-attention mechanism aligns clinical metadata with spatial image features during prediction.

**Figure 4:**
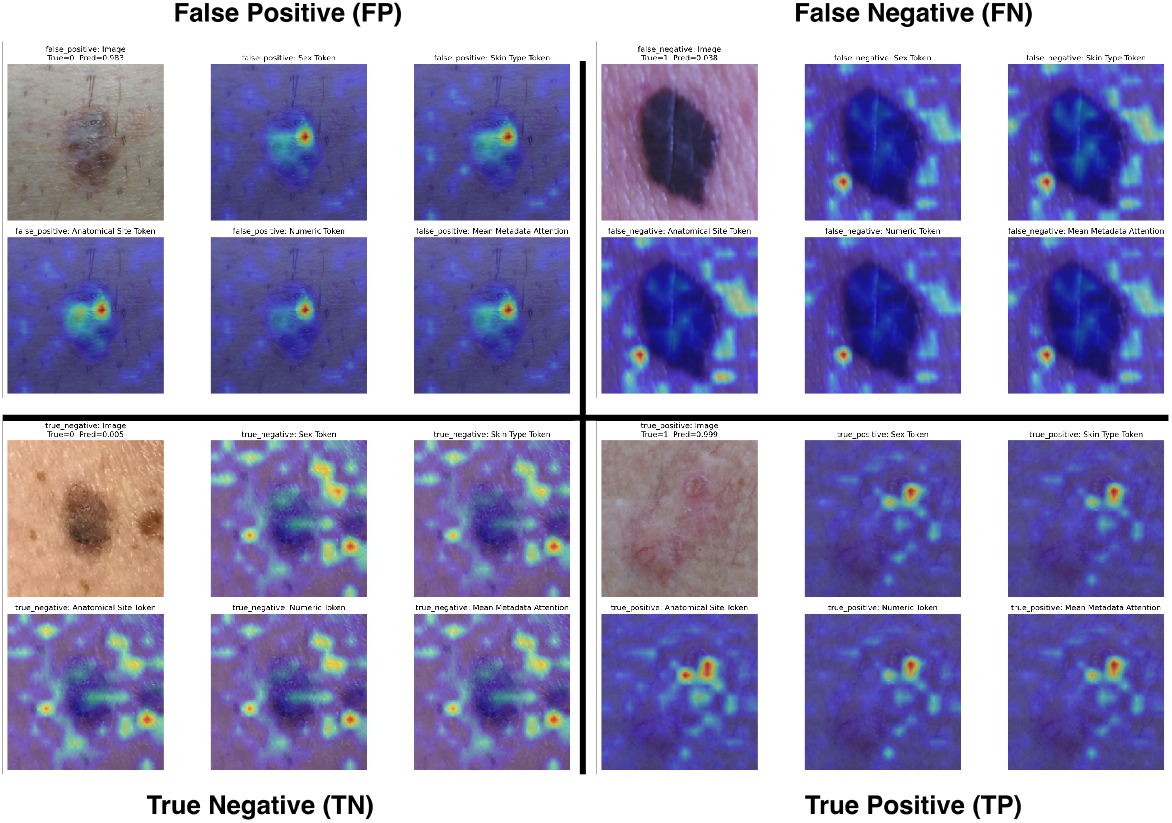
Qualitative visualization of cross-attention maps conditioned on different metadata tokens. Correct predictions focus attention on clinically relevant lesion structures, whereas misclassified samples exhibit more diffuse or misplaced attention patterns.

#### Clinical Feature Importance

Permutation analysis was used to assess the contribution of each metadata variable (Table 3). Removing all metadata produced the largest performance decline (AUC drop of 0.0453), confirming that patient context provides important complementary information beyond image features alone. Among individual variables, sex and Fitzpatrick skin type showed the largest performance reductions when permuted, indicating strong contributions to the model’s predictions. Age and lesion diameter produced moderate declines, whereas anatomical site had minimal impact on performance in this dataset.

**Table 3:** Clinical Feature Importance via Permutation Analysis. Performance changes when permuting each metadata feature across the test set while keeping the trained multimodal model fixed. Negative values indicate performance degradation relative to the full model.

| Configuration | AUC | AUPRC |
| --- | --- | --- |
| Full model (all metadata) | 0.9818 | 0.9924 |
| Permute anatomical site | 0.9826 (+0.0008) | 0.9930 (+0.0006) |
| Permute sex | 0.9707 (-0.0111) | 0.9888 (-0.0036) |
| Permute lesion diameter | 0.9784 (-0.0034) | 0.9911 (-0.0013) |
| Permute Fitzpatrick skin type | 0.9711 (-0.0106) | 0.9889 (-0.0035) |
| Permute age | 0.9768 (-0.0049) | 0.9890 (-0.0034) |
| Image only (no metadata) | 0.9365 (-0.0453) | 0.9790 (-0.0134) |

## Discussion

This study demonstrates that the manner in which clinical metadata is integrated with image features critically influences the performance of multimodal diagnostic systems. While the image-only model already achieved strong discrimination (AUC = 0.9776), simple late fusion of metadata slightly degraded performance, indicating that naive concatenation may introduce noise rather than meaningful clinical context. In contrast, the cross-attention architecture produced the best overall performance (AUC = 0.9818, AUPRC = 0.9924) and improved calibration (ECE = 0.0379; Brier score = 0.0323). These results suggest that enabling structured interaction between modalities allows patient information to meaningfully influence visual representation learning.

The permutation-based analysis further illustrates the importance of clinical context. Removing all metadata produced the largest performance decline (AUC drop of 0.0453), confirming that patient information provides complementary signal beyond image features alone. Among individual variables, sex and Fitzpatrick skin type produced the largest performance reductions when permuted, while lesion diameter and age showed moderate contributions. These findings suggest that contextual patient characteristics can meaningfully guide visual interpretation, consistent with how dermatologists incorporate demographic and lesion characteristics when assessing malignancy risk.

Despite these improvements, the performance gain over the strong image-only baseline was modest and not statistically significant in bootstrap comparisons. This likely reflects the relatively small size of the dataset and the already high predictive signal present in dermoscopic images. Larger and more diverse datasets may be necessary to fully realize the potential of multimodal reasoning in clinical AI systems.

Several limitations should be noted. First, the study relies on a single dataset (PAD-UFES-20), and external validation across institutions and imaging conditions will be necessary to establish generalizability. Second, the dataset contains an enriched proportion of malignant lesions relative to real-world screening populations, which may influence model calibration and decision thresholds. Third, only a limited set of structured metadata variables were included; realworld clinical reasoning often incorporates additional contextual information such as patient history, lesion evolution, and longitudinal health records.

Overall, our findings highlight that incorporating clinical metadata can improve both predictive accuracy and calibration, but only when the integration mechanism allows meaningful interaction between modalities. Attention-based multimodal architectures provide a principled framework for modeling this interaction and represent a promising direction for developing clinically useful decision-support systems in dermatology.

## Data Availability

All data produced in the present work are contained in the manuscript

## Code Availability

https://github.com/krishna-mridhacase/multimodal-skin-lesion-cross-attention

## Notes

### Competing Interest Statement

The authors have declared no competing interest.

### Funding Statement

This study did not receive any funding

### Author Declarations

The study used the publicly available PAD-UFES-20 dataset, which contains dermoscopic images and associated clinical metadata. The dataset includes individual-level observations; however, all patient information provided in the dataset is fully de-identified. No personally identifiable information is included. All data used in the study were obtained from this publicly released dataset, and no additional patient-level data were collected by the authors.

## References

1. Lee KJ, Betz-Stablein B, Stark MS, Janda M, McInerney-Leo AM, Caffery LJ, et al. The Future of Precision Prevention for Advanced Melanoma. Frontiers in Medicine. 2022;Volume 8 - 2021.

2. Esteva A, Kuprel B, Novoa RA, Ko J, Swetter SM, Blau HM, et al. Dermatologist-level classification of skin cancer with deep neural networks. Nature. 2017;542(7639):115–8.

3. Haenssle HA, Fink C, Schneiderbauer R, Toberer F, Buhl T, Blum A, et al. Man against machine: diagnostic performance of a deep learning convolutional neural network for dermoscopic melanoma recognition in comparison to 58 dermatologists. Annals of Oncology. 2018;29(8):1836–42.

4. Codella NC, Gutman D, Celebi ME, Helba B, Marchetti MA, Dusza SW, et al. Skin lesion analysis toward melanoma detection: A challenge at the 2017 ISBI, hosted by the ISIC. In: 2018 IEEE 15th International Symposium on Biomedical Imaging (ISBI); 2018. p. 168–72.

5. Tschandl P, Rinner C, Apalla Z, Argenziano G, Codella N, Halpern A, et al. Human–computer collaboration for skin cancer recognition. Nature Medicine. 2020;26(8):1229–34.

6. He K, Zhang X, Ren S, Sun J. Deep residual learning for image recognition. In: Proceedings of the IEEE Conference on Computer Vision and Pattern Recognition (CVPR); 2016. p. 770–8.

7. Dosovitskiy A, Beyer L, Kolesnikov A, Weissenborn D, Zhai X, Unterthiner T, et al. An Image is Worth 16×16 Words: Transformers for Image Recognition at Scale. In: International Conference on Learning Representations (ICLR); 2021.

8. Daneshjou R, Smith MP, Sun MD, Rotemberg V, Zou J. Disparities in dermatology AI performance on a diverse, curated clinical image set. Science Advances. 2022;8(32):eabq6147.

9. Huang SC, Pareek A, Seyyedi S, Banerjee I, Lungren MP. Fusion of medical imaging and electronic health records using deep learning: a systematic review and implementation guidelines. NPJ Digital Medicine. 2020;3(1):136.

10. Yan K, Peng Y, Sandfort V, Bagheri M, Lu Z, Summers RM. Multi-task learning for chest X-ray abnormality classification on noisy labels. In: Medical Imaging 2018: Computer-Aided Diagnosis. vol. 10575; 2018. p. 105751G.

11. Zhang C, Yang Z, He X, Deng L. Multimodal intelligence: representation learning, information fusion, and applications. IEEE Journal of Selected Topics in Signal Processing. 2020;14(3):478–93.

12. Guan Y, Zhang J, Tian K, Yang S, Dong P, Xing L, et al. Multi-modal learning for predicting the genotype of clear cell renal cell carcinoma. In: IEEE International Symposium on Biomedical Imaging (ISBI); 2022. p. 1–5.

13. Lu J, Batra D, Parikh D, Lee S. ViLBERT: Pretraining task-agnostic visiolinguistic representations for vision-and-language tasks. In: Advances in Neural Information Processing Systems (NeurIPS). vol. 32; 2019.

14. Huang SC, Shen L, Lungren MP, Yeung S. GLORIA: A multimodal global-local representation learning framework for label-efficient medical image recognition. In: Proceedings of the IEEE/CVF International Conference on Computer Vision (ICCV); 2021. p. 3942–51.

15. Müller D, Soto-Rey I, Kramer F. Joint learning of histopathological image and genomic data for improved survival prediction in glioma. IEEE Journal of Biomedical and Health Informatics. 2021;25(2):497–507.

16. Bahdanau D, Cho K, Bengio Y. Neural machine translation by jointly learning to align and translate. arXiv preprint arXiv:14090473. 2014.

17. Vaswani A, Shazeer N, Parmar N, Uszkoreit J, Jones L, Gomez AN, et al. Attention Is All You Need. In: Advances in Neural Information Processing Systems (NeurIPS). vol. 30; 2017.

18. Pacheco AGC, Krohling RA, Barata C, et al. PAD-UFES-20: A skin lesion dataset composed of patient data and clinical images collected from smartphones. Data in Brief. 2020;32:106221.

